# Neighborhood income and physical distancing during the COVID-19 pandemic in the U.S.

**DOI:** 10.1101/2020.06.25.20139915

**Authors:** Jonathan Jay, Jacob Bor, Elaine Nsoesie, Sarah Ketchen Lipson, David K. Jones, Sandro Galea, Julia Raifman

## Abstract

**Introduction:** Although physical distancing has been the primary strategy to reduce the spread of COVID-19 in the U.S., people’s ability to distance may vary by socioeconomic characteristics, leading to higher transmission risk in low-income neighborhoods.

**Methods:** We used mobility data from a large, anonymized sample of smartphone users to assess the relationship between neighborhood median household income and physical distancing during the COVID-19 epidemic. We assessed changes in several behaviors including: spending the day entirely at home; working outside the home; and visits to supermarkets, parks, hospitals, and other locations. We also assessed differences in effects of state policies on physical distancing across neighborhood income levels.

**Results:** We found a strong gradient between neighborhood income and physical distancing. Compared to January and February 2020, the proportion of individuals spending the day entirely at home in April 2020 increased by 10.9 percentage points in low-income neighborhoods and by 27.1 percentage points in high-income neighborhoods. During April 2020, people in low-income neighborhoods were more likely to work outside the home, compared to people in higher-income neighborhoods, but not more likely to visit non-work locations. State physical distancing orders were associated with a 1.5 percentage-point increase (95% CI [0.9, 2.1], *p* < 0.001) in staying home in low-income neighborhoods and a 2.4 percentage point increase (95% CI [1.4, 3.4], *p* < 0.001) in high-income neighborhoods.

**Discussion:** People in lower-income neighborhoods have faced barriers to physical distancing, particularly the need to work outside the home. State physical distancing policies have not mitigated these disparities.

## Introduction

Physical distancing has been the primary strategy to limit the spread of COVID-19 in the United States. Physical distancing (also called “social distancing”) entails reducing contacts between non-household members in order to reduce opportunities for transmission from infected to susceptible individuals. To promote physical distancing, most U.S. states closed schools, mandated business closures, and issued “stay-at-home” orders directing residents to avoid unnecessary trips. These measures have been essential to prevent worst-case scenarios involving millions of deaths.^1,2^

Although there is evidence that people stayed home and that new cases of COVID-19 declined,^1^ evidence suggests unequal declines in the burden of COVID-19. While case data disaggregated by income are not available, COVID-19 case and death rates have risen fastest in low-income communities.^3,4^ An association between lower neighborhood income and COVID-19 risk is also consistent with data showing higher COVID-19 mortality among racial and ethnic minorities,^5^ whose socioeconomic position is systematically lower, on average, than that of white Americans, and who disproportionately reside in low-income neighborhoods due to a long history of discriminatory housing policy.^6,7^

Financial constraints to physical distancing may have been an important factor contributing to higher COVID-19 burden among economically marginalized populations.^4^ At businesses that have remained open during the pandemic, low-income workers have reported less ability to work from home relative to higher wage-earners.^8^ At these workplaces, most workers were not eligible for unemployment insurance unless they could document a COVID-19 diagnosis or exposure.^9^ Although many states began closing businesses and ordering residents to stay home in the second half of March, businesses deemed essential remained open, and staffed predominantly by low-wage workers.^10,11^ It was not until mid-April that some states began requiring people to wear masks in public spaces to reduce COVID-19 transmission, and many states still have not done so.^9^ In this context, low-income workers have had to choose between staying home and losing their income or going to work and risking exposure to COVID-19 for themselves and their households and neighbors. Given that those in low-income households typically have little savings,^12^ losing income could bring other health and safety risks, including homelessness and food insecurity.

In this article, we test two main hypotheses. First, we hypothesized that residents of low-income neighborhoods were less likely than residents of higher-income neighborhoods to stay home in response to COVID-19 (Hypothesis 1). This difference was driven by work-related demands (Hypothesis 1a) and not by visits to places other than work (Hypothesis 1b). Second, we hypothesized that state policies that ordered non-essential businesses to close, and for residents to stay at home, increased the gap in physical distancing between low-income and high-income neighborhoods (Hypothesis 2).

To test these hypotheses, we use longitudinal mobility data derived from smartphones during the first four months of the COVID-19 epidemic in the United States. We focus our analyses at the neighborhood level because community-level physical distancing is thought to be a key driver of disparities in exposure to COVID-19. It is also the smallest available unit of geography for which de-identified mobility data are available. To our knowledge, this is the first national study of the relationship between neighborhood income and physical distancing behaviors.

## Methods

### Data

#### Mobility metrics

We obtained mobility data from SafeGraph, a data company that aggregates anonymized location data from smartphone applications. Several other studies have used SafeGraph data to examine U.S. mobility during the COVID-19 pandemic.^13–17^ We used data derived from an average sample of approximately 19 million smartphone devices observed per day for 50 states and the District of Columbia. We included observations from January 6, 2020 through May 1, 2020, excluding one date in February known to contain measurement errors. For physical distancing behaviors, SafeGraph aggregated these data for each calendar date at the U.S. census block group (BG) level. BGs are smaller than census tracts and typically contain 600-3,000 people. There are 217,740 BGs in the U.S., 99.6% of which were included in the SafeGraph data.

We did not expect the SafeGraph sample to be representative of the general population, because smartphone ownership varies across socio-demographic characteristics, particularly age and income.^18^ However, we found only a small, positive correlation (Pearson’s *r* = 0.02) between the device-to-population ratio and BG median household income. For linear regressions, we weight models according to the total devices observed for each unit of analysis.

#### Staying at home

Our primary outcome of interest was the proportion of smartphone users who spent all day at home, for each date. SafeGraph inferred a smartphone user’s home location (a 152m by 152m cell) based on where their device was located overnight for most nights during the previous 6 weeks. A smartphone user was considered to be at home all day when their device was observed within the inferred home location and nowhere else on a given date.

#### Working outside the home

For a secondary analysis, our outcome was the proportion of smartphone users who were inferred to have gone to work outside the home on a given day. The numerator included smartphone users whose behavior was consistent with full-time or part-time work (stopping at a location for at least 3 hours between 8am and 6pm) or delivery work (stopping at 4 or more locations for less than 20 minutes each). While this metric does not count overnight work shifts and likely undercounts overall work (as suggested by relatively low overall proportions of smartphone users recorded as working— approximately 20-25%/day at baseline, whereas labor force participation among U.S. adults is typically over 60%^19^), our primary interest was temporal trends in work outside the home, which it appeared adequate to detect. For instance, this metric recorded large reductions in working on national holidays in January and February 2020.

#### Visits to non-work locations

We used different SafeGraph datasets to measure trends in *non-work visits* to seven categories of places: parks and playgrounds, hospitals, supermarkets, carryout restaurants, places of worship, convenience stores, and liquor stores. These data were obtained at the point of interest (“POI”) level (*e*.*g*., one record for each of the 50,760 supermarkets in our sample, see **Table 1**) and omit visits from individuals whose workplace is the point of interest. We only included locations in metropolitan counties, population 100,000 and above, to improve comparability.

**Table 1:** Sample characteristics.

| Income quintile | Income range (USD) | Census block groups |  |  |  | Points of interest |  |  |  |  |  |  |
| --- | --- | --- | --- | --- | --- | --- | --- | --- | --- | --- | --- | --- |
|  |  | Sample devices |  | Population |  | Beer, wine & liquor stores | Carryout | Convenience stores | Hospitals | Parks & playgrounds | Places of worship | Supermarkets |
|  |  | Mean | SD | Mean | SD | *n* |  |  |  |  |  |  |
| 1 | Up to 40870 | 68 | 47 | 1265 | 701 | 3135 | 11612 | 2764 | 287 | 9203 | 17759 | 10551 |
| 2 | 40870-54305 | 83 | 61 | 1423 | 819 | 3058 | 21531 | 4289 | 706 | 9952 | 14267 | 10980 |
| 3 | 54306-69769 | 93 | 77 | 1527 | 952 | 3397 | 28401 | 4978 | 1123 | 12626 | 14840 | 10993 |
| 4 | 69770-93749 | 104 | 99 | 1679 | 1134 | 3866 | 28927 | 4511 | 935 | 15236 | 14677 | 10289 |
| 5 | 93750 and above | 106 | 120 | 1737 | 1271 | 3473 | 22580 | 2934 | 597 | 17285 | 11331 | 7947 |
| **Full sample** |  | 88 | 83 | 1483 | 990 | 16929 | 113051 | 19476 | 3648 | 64302 | 72874 | 50760 |

*Notes.* Census block groups (BGs) were the units of analysis for changes in the proportion of smartphone users staying home all day and going to work outside the home, for January 6 through May 1, 2020. BGs were assigned income quintiles based on the population-weighted quintiles for median household income for all U.S. BGs in 2018. Points of interest (POIs) were the units of analysis for changes in non-work visits occurring between March 1 and May 2, 2020. POIs were assigned income quintiles based on the typical visitor to each POI in January and February 2020. For more details, see Methods.

Broadly, our approach was to assign each POI an income quintile corresponding to the household income level of the neighborhood where the POI’s typical visitor resides, then track total visits to each POI during the COVID-19 period. More specifically, we assigned POI visitor income based on the median BG income of visitors to each POI during January and February 2020, weighted by the number of visits from each BG. We adjusted those BG-level visit counts to account for variation in the device-to-population ratio for each BG. Next, we aggregated visitor counts from January 6 to May 3 by POI and week. We also adjusted these visits to account for variation in device-to-population ratio.

#### State-level policies

We used a publicly available database of state-level COVID response policies, including physical distancing measures.^9^ These data were collected by tracking news coverage and verifying news reports against government websites. Policy measures instituted prior to May 1, 2020 were included in the analysis. For this analysis, we analyzed the effects of a physical distancing policy indicator that combined (a) non-essential business closures and (b) “stay-at-home” orders (also called “shelter-in-place” orders). According to our measure, the policy exposure began as soon as either measure went into effect. We used this indicator for three reasons. First, business closures and stay-at-home orders were not always readily distinguishable from one another. For example, although Connecticut and Kentucky adopted measures that officials referred to as stay-at-home orders, these orders did not mandate staying at home, but did require the closure of non-essential businesses. Second, many states adopted both of these measures, either at the same time or in close temporal proximity, creating strong collinearity in exposure. Third, both of these measures aim to address COVID-19 risk by reducing potential exposures outside the home, except for workers deemed essential.

On March 19, California issued a stay-at-home order and closed non-essential businesses, while Pennsylvania also closed non-essential businesses. By May 1^st^, 45 states and the District of Columbia had issued stay-at-home orders and/or non-essential business closures. 11 states had implemented, and then lifted, at least one of these measures. Several of these states ceased business closures while keeping stay-at-home orders in place. Since the impact of these partial re-openings was unknown, to be conservative in our estimation of policy effects we coded these states as having no policy exposure starting on the date when either measure was lifted.

#### Other variables

For each BG in the U.S. for which SafeGraph data were available (n = 177,452), we obtained 2018 American Community Survey data on median household income and used these data to calculate the population-weighted income quintile for each BG. These quintiles ranged from median household incomes of $40,870 and below to $93,750 and above. See **Table 1**. Urbanicity was based on county-level classifications from the National Center for Health Statistics,^20^ and we used state-level classifications from the U.S. Census Bureau to assign regions.

We obtained daily COVID-19 case counts by state from a repository compiled by the New York Times from state government sources.^21^ We anticipated that differences in the severity of local COVID-19 outbreaks could influence residents’ decision to stay at home, notwithstanding state policies. This association could confound the association between physical distancing policies and staying at home, particularly since some of the first states to issue restrictions were states with early outbreaks (*e*.*g*., California, New York, Massachusetts). Therefore, we included case counts as covariates in regression models described below.

### Analysis

#### Changes in mobility by neighborhood income

Physical distancing behaviors, as measured by the proportion of smartphone users staying home all day, was our primary outcome of interest. For the proportion of smartphone users staying at home, we aggregated daily BG-level data by week and income quintile to assess level and trends over time. We examined these dynamics further by disaggregating by urbanicity and by region. Using the daily BG-level data, we estimated changes for each income quintile relative to baseline, comparing a “pre” period (January 6-February 29, 2020) with a “post” period (April 1-April 30, 2020) in OLS regression models.

To assess how work contributed to physical distancing, we conducted similar analyses of trends in the proportion of smartphone users working outside the home. To assess the role of non-work activities outside the home, we calculated changes in visits within each visitor income level and POI category over similar pre (January 6-Mar 1) and post (Apr May 3) periods. We normalized these visit counts against pre-period means and report changes as the proportion of pre-period visits that occurred during the post-period. For places of worship, we also conducted separate exploratory analysis of weekday and weekend visits, to assess whether these spaces were being used for religious services or for other functions.

#### Effects of state physical distancing orders on mobility by neighborhood income

To estimate the effects of state physical distancing orders, we used a difference-in-differences (DiD) linear regression model with two-way fixed effects for every state and date. Fixed effects by state account for each state’s time-invariant characteristics, while fixed effects by date account for time-variant but state-invariant characteristics.^22^ The treatment variable was a binary indicator set to 1 for each date in a given state when physical distancing orders were in effect, and otherwise set to 0. When states revoked physical distancing orders, the indicator reverted to 0. To control for differences in the severity of local COVID-19 outbreaks, we further adjusted for the count of cases in the state, with a one-day time lag. We included dummies for one-day lagged cases, with cut points at case counts of 1, 10, 100, 1000, and 10000 and above, with 0 as the reference group. This approach minimized assumptions about the functional form of the relationship between cases and mobility, although using log-normalized case counts instead produced similar results (available upon request).

To examine the differential effect of physical distancing orders across income levels, we interacted the income quintile indicator with every other covariate (*i*.*e*., the model was *fully interacted*). The regression coefficients of interest were those corresponding to treatment in the lowest-income quintile (the reference level) and the interaction terms *treatment* * *income quintile* for every other income quintile. These interaction terms estimated the difference between the treatment effect at each income quintile and the lowest-income quintile.

We also estimated event study models to assess trends in mobility in the days before and after states instituted physical distancing orders. This approach allows for testing the DiD model assumption that intervention and non-intervention groups had parallel pre-intervention time trends, as well as to examine temporal heterogeneity in policy effects. In the event study models, we replaced the binary policy indicator with binary indicators for living in intervention states in a series of one-day periods up to 14 days before and after policy changes. The reference group was being in a comparison state or being in an intervention state on the day before policy enactment. We estimated these models separately for each income stratum and omitted interaction terms.

All DiD and event study models were weighted by daily device counts to account for the greater precision provided by observations based on more users. We also clustered the models’ standard errors by state to account for serial autocorrelation. However, since cluster-robust standard errors may not be reliable in DiD analyses with small samples,^23^ we conducted placebo tests to validate statistical significance in the main model. In these tests, we re-estimated the model with the policy exposure randomly reassigned across states, such that any estimated association was necessarily spurious. We then compared the *t-*statistic of our original finding to those observed over 1000 iterations of the placebo treatment to calculate an alternative *p* value.

We used OLS regression for all models. Although our outcome variable (the proportion of smartphone users staying home all day) was bounded [0, 1], OLS was an acceptable approach because very few observations approached these limits.^24^ Poisson and beta regressions yielded similar results (available upon request).

Analyses were conducted in R software. Since the mobility data were anonymized and other data were publicly available, this study was exempted from institutional review board review as non-human subjects research.

## Results

### Days spent entirely at home

We found an increase in physical distancing for all income levels from January-February 2020 to April 2020. Before these changes, people residing in higher-income neighborhoods stayed home the least, while people residing in lower-income neighborhoods stayed home the most. Afterwards, this relationship inverted. **Figure 1**.

**Figure 1.**
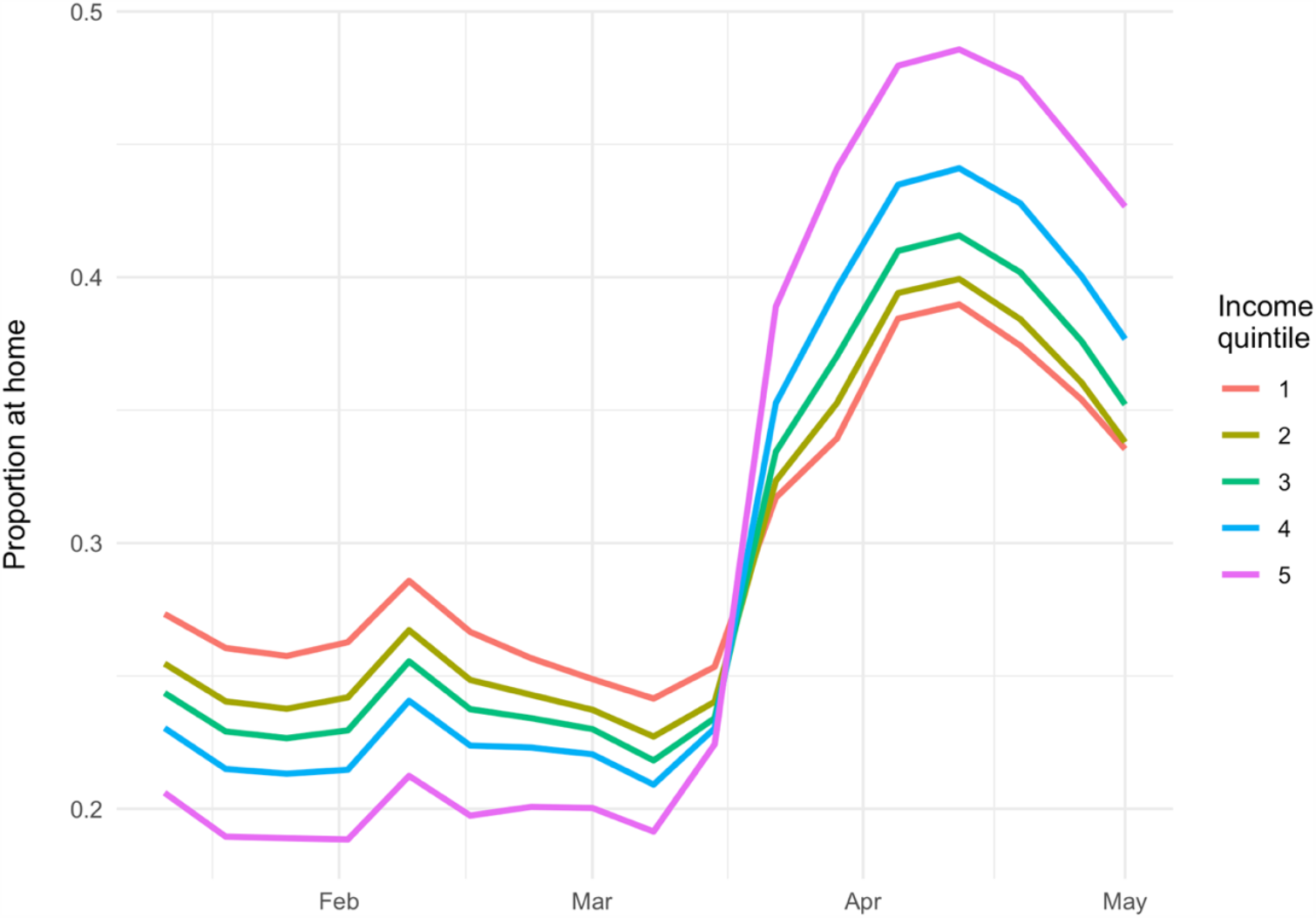
Proportion of smartphone users staying home all day. *Notes*: Income quintile 1 represents the lowest-income group. Outcomes are presented as weekly averages. Period covered is January 6, 2020, through May 1, 2020.

In the lowest-income neighborhoods, staying home increased by 10.9 percentage points during the physical distancing period. In the highest-income neighborhoods, staying home increased by 27.1 percentage points during the physical distancing period. This increase in the highest-income neighborhoods was 16.2 percentage points greater (95% CI [16.2, 16.3], *p* < 0.001) than the increase observed in the lowest-income neighborhoods. **Table 2**.

**Table 2:** Proportion of smartphone users spending entire day at home.

| Neighborhood income quintile | Mean |  | Absolute difference (95% CI) | Difference relative to Q1 (95% CI) |
| --- | --- | --- | --- | --- |
|  | January-February 2020 | April 2020 |  |  |
| Q1 (lowest) | 26.3 | 37.2 | 10.9<br>(10.8, 10.9) | -- |
| Q2 | 24.5 | 38.0 | 13.5<br>(13.4, 13.5) | 2.6<br>(2.6, 2.6) |
| Q3 | 23.5 | 39.6 | 16.2<br>(16.1, 16.2) | 5.3<br>(5.3, 5.3) |
| Q4 | 22.1 | 42.2 | 20.0<br>(19.9, 20.1) | 9.1<br>(9.1, 9.2) |
| Q5 (highest) | 19.7 | 46.8 | 27.1<br>(27.0, 27.2) | 16.2<br>(16.2, 16.3) |

*Notes.* All $p$ values < 0.001. To calculate differences from “pre” (January-February 2020) to “post” (April 2020) COVID-19 related changes, we used OLS regressions interacting a “post” period indicator with income quintile, using quintile 1 as the reference group.

Levels of physical distancing and disparities by neighborhood income were greater in larger cities and more urbanized areas. However, the gradient in physical distancing by neighborhood income was apparent even in rural areas. The highest level of physical distancing for any single group was among the highest-income quintile living in the suburbs of large cities (“fringe metro” areas). **Figure 2**.

**Figure 2.**
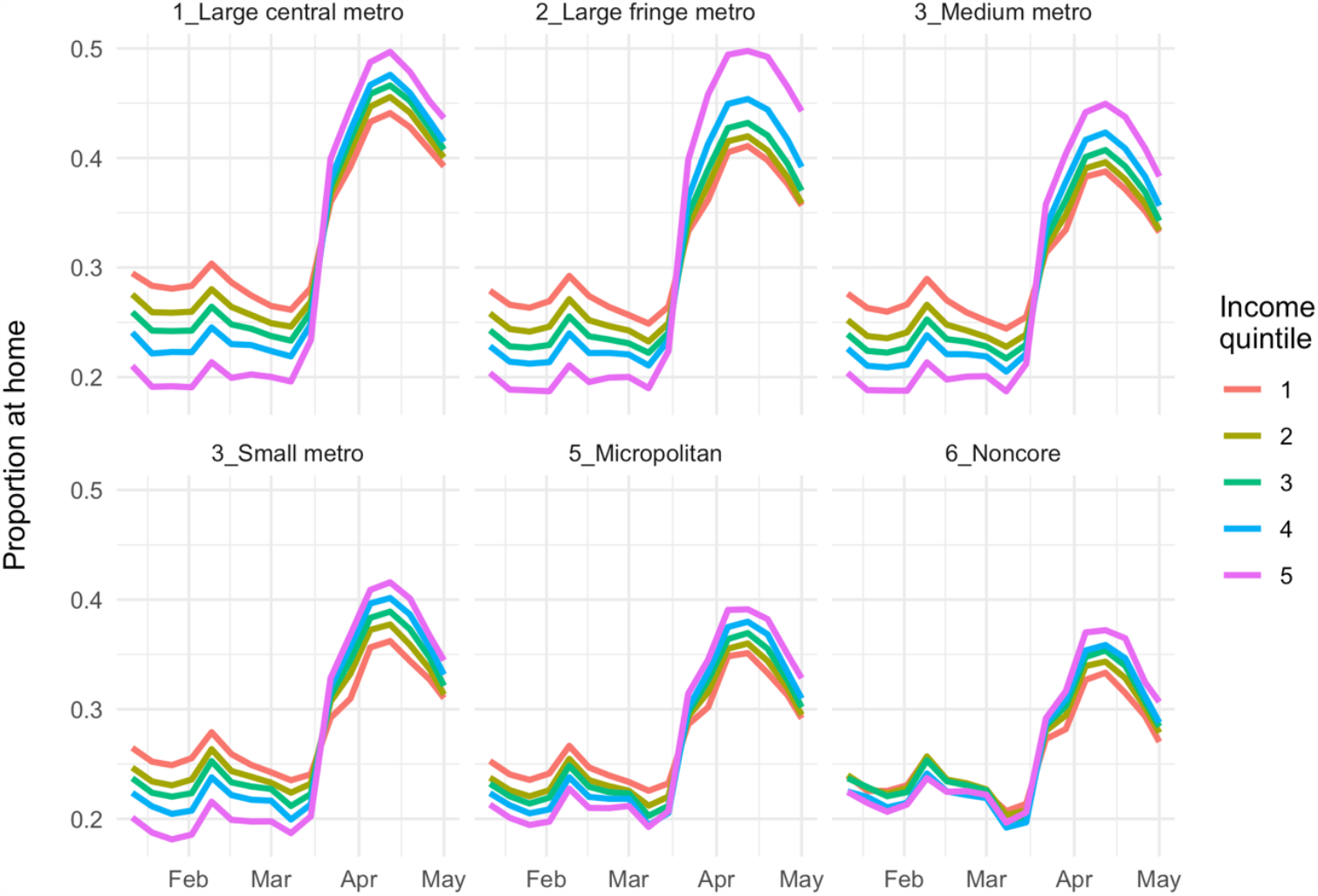
Proportion of smartphone users staying home all day by level of urbanicity. *Notes*: Income quintile 1 represents the lowest-income group. Outcomes are presented as weekly averages. Period covered is January 6, 2020, through May 1, 2020. Levels of urbanicity are National Center for Health Statistics classifications.^20^

Physical distancing rates were highest in the Northeast and lowest in the South. Disparities in distancing by neighborhood income were similar in all regions. **Figure S1**.

**Fig. S1.**
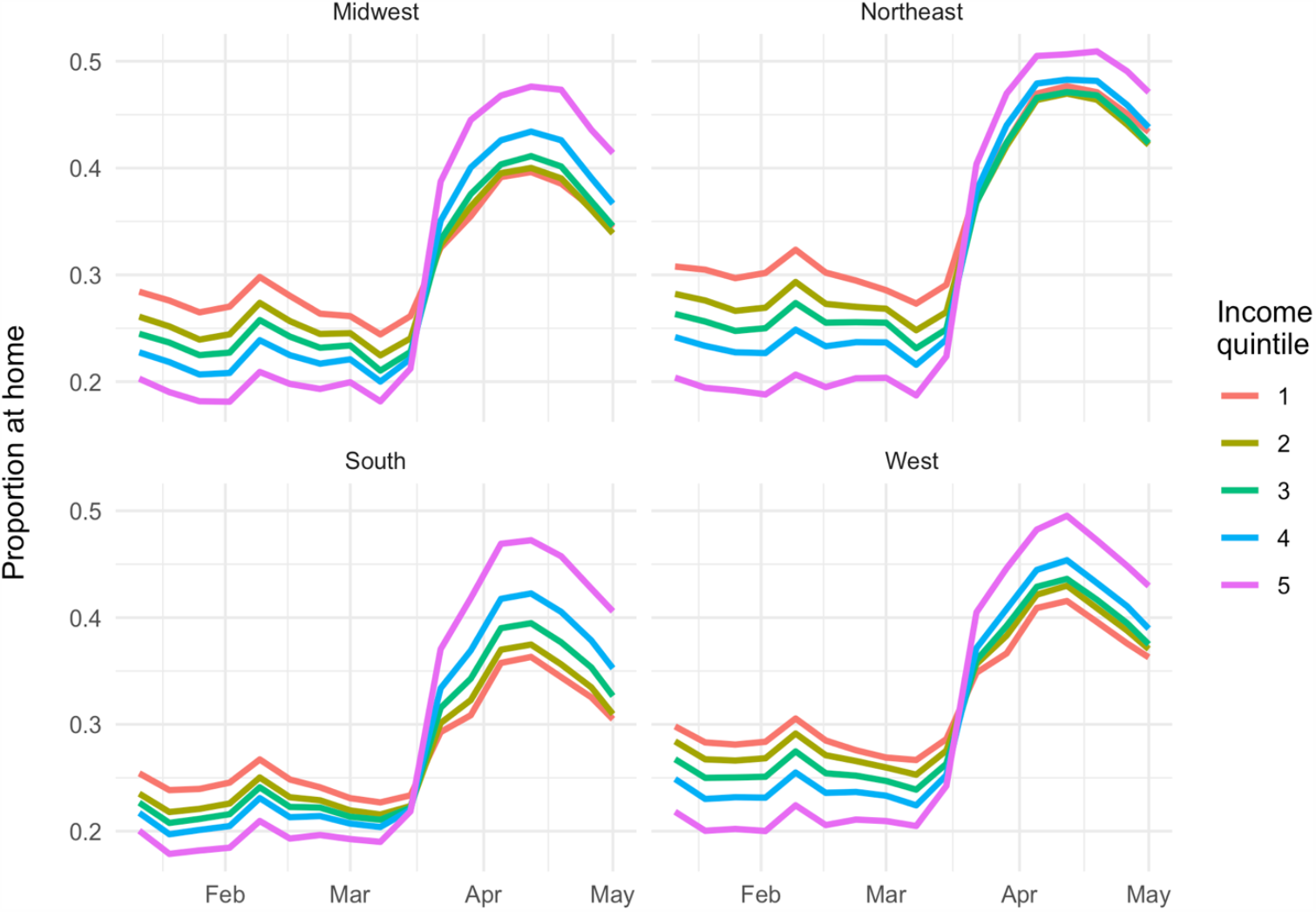
Proportion of smartphone users staying home all day by region. *Notes*: Income quintile 1 represents the lowest-income group. Outcomes are presented as weekly averages. Period covered is January 6, 2020, through May 1, 2020. Regions are U.S. Census Bureau classifications.

### Days working outside the home

For each neighborhood income quintile, we found reductions in working outside the home that corresponded with increases in physical distancing. **Figure 3**.

**Figure 3:**
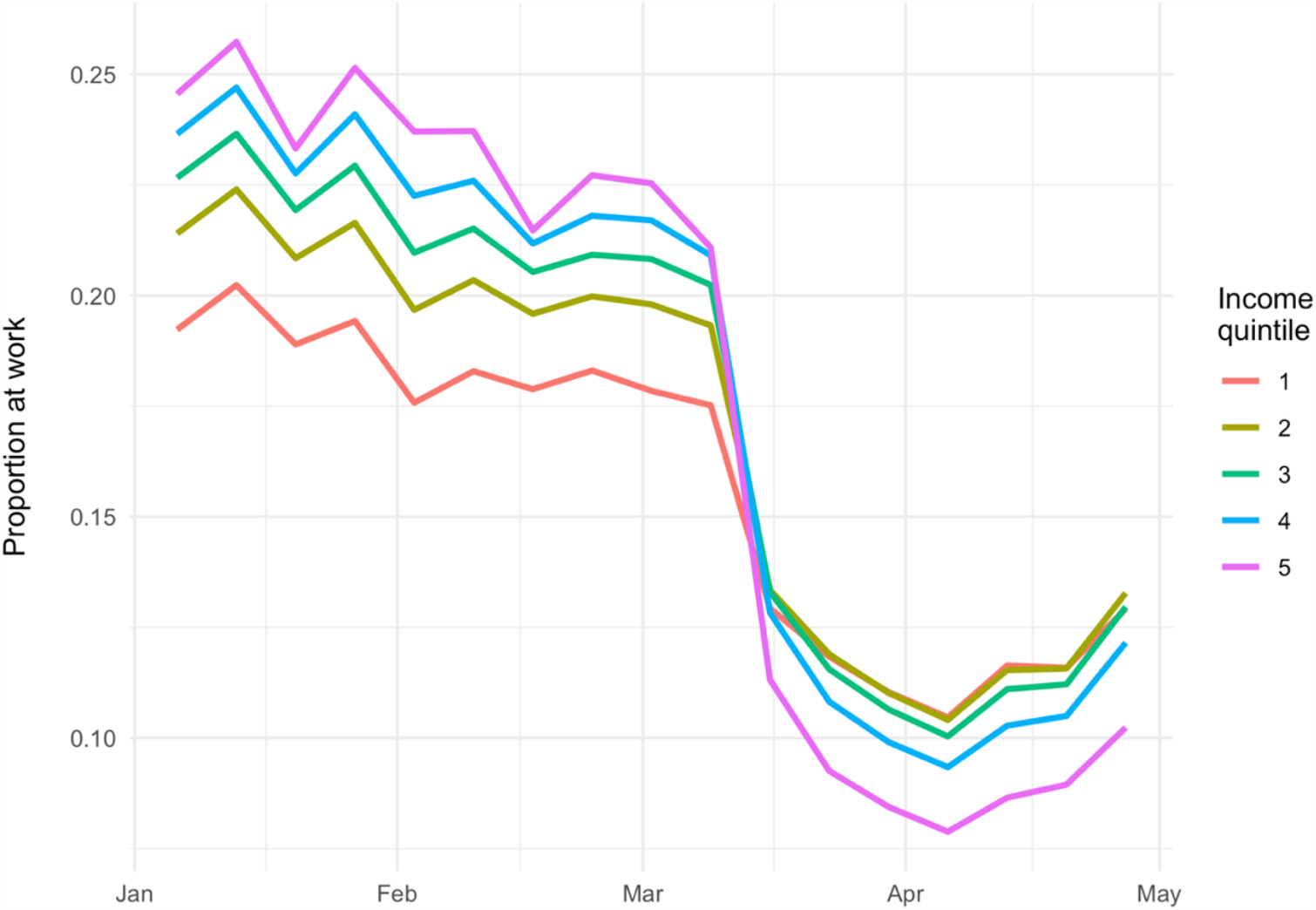
Proportion of smartphone users working outside the home. *Notes:* Income quintile 1 represents the lowest-income group. Outcomes are presented as weekly averages. Period covered is January 6, 2020, through May 1, 2020.

The highest-income group worked outside the most at baseline and the least during COVID-19. Reductions in working outside the home were largest among the highest-income group, which reduced days at work by 15.5 percentage points. This reduction was 7.9 percentage points greater (95% CI [7.9, 7.9], *p* < 0.001) than the reduction in the lowest-income group, which reduced days at work by 7.6 percentage points. **Table 3**.

**Table 3:** Percentage of smartphone users working outside the home.

| Neighborhood income quintile | Mean |  | Absolute difference (95% CI) | Difference relative to Q1 (95% CI) |
| --- | --- | --- | --- | --- |
|  | January-February 2020 | April 2020 |  |  |
| Q1 (lowest) | 18.9 | 11.3 | -7.6<br>(-7.6, -7.6) | -- |
| Q2 | 20.9 | 11.3 | -9.7<br>(-9.7, -9.6) | -2.1<br>(-2.1, -2.0) |
| Q3 | 22.1 | 10.9 | -11.2<br>(-11.3, -11.2) | -3.6<br>(-3.7, -3.6) |
| Q4 | 23.1 | 10.1 | -13.0<br>(-13.0, -13.0) | -5.4<br>(-5.4, -5.4) |
| Q5 (highest) | 24.1 | 8.6 | -15.5<br>(-15.5, -15.5) | -7.9<br>(-7.9, -7.9) |

*Notes.* All $p$ values < 0.001. To calculate differences from “pre” (January-February 2020) to “post” (April 2020) COVID-19 related changes, we used OLS regressions interacting a “post” period indicator with income quintile, using quintile 1 as the reference group.

### Non-work activities outside the home

Over the same period, visits declined to all categories of non-work locations across all income levels. **Table 4, Figure 4**.

**Table 4:** Non-work activities outside the home: percent change in visits by category.

| Neighborhood income quintile of typical visitor | Percent change in visits (95% CI) |  |  |  |  |  |  |
| --- | --- | --- | --- | --- | --- | --- | --- |
|  | Beer, wine and liquor stores | Carryout restaurants | Convenience stores | Hospitals | Parks & playgrounds | Places of worship | Supermarkets |
| Q1 (lowest) | -44.6<br>(-48.4, -40.9) | -57.6<br>(-59.5, -55.8) | -43.2<br>(-45.4, -40.9) | -57.5<br>(-67.8, -47.1) | -58.5<br>(-60.8, -56.1) | -59.2<br>(-60.5, -58.0) | -39.0<br>(-40.7, -37.3) |
| Q2 | -39.8<br>(-42.2, -37.4) | -37.0<br>(-37.9, -36.1) | -37.5<br>(-39.3, -35.8) | -54.0<br>(-58.9, -49.1) | -53.2<br>(-55.3, -51.1) | -64.0<br>(-65.1, -62.9) | -28.8<br>(-30.2, -27.3) |
| Q3 | -38.0<br>(-40.4, -35.6) | -37.6<br>(-38.4, -36.8) | -37.8<br>(-39.4, -36.2) | -55.8<br>(-59.9, -51.7) | -53.7<br>(-55.9, -51.5) | -72.8<br>(-75.2, -70.5) | -27.2<br>(-28.7, -25.7) |
| Q4 | -42.6<br>(-46.0, -39.3) | -44.0<br>(-44.9, -43.0) | -40.9<br>(-42.6, -39.3) | -58.0<br>(-63.1, -53.0) | -59.0<br>(-61.2, -56.8) | -80.7<br>(-82.2, -79.2) | -29.1<br>(-30.6, -27.7) |
| Q5 (highest) | -47.4<br>(-50.0, -44.7) | -54.4<br>(-55.3, -53.5) | -48.6<br>(-50.7, -46.6) | -57.8<br>(-63.7, -51.9) | -62.0<br>(-63.7, -60.2) | -86.6<br>(-88.3, -84.9) | -33.6<br>(-35.1, -32.0) |

*Notes.* All $p$ values < 0.001. To calculate differences from “pre” (January 6-Mar 1, 2020) to “post” (April 6-Mar 3, 2020) COVID-19 related changes, we used OLS regressions estimating “post” effects, stratified within each location category and visitor income quintile. Values were normalized against the pre-period mean within each category and income quintile prior to modeling, to estimate proportional changes in visits from pre to post. These are reported here as percentages. For comparability, metrics were calculated for locations in urban counties (population 100,000 and above) only.

**Table 5:** Difference-in-differences (DiD) linear regression estimates: effects of physical distancing orders on staying home all day.

| Neighborhood income quintile | DiD Estimate | 95% CI | P value | Difference in DiD Estimate<br>relative to Q1 | 95% CI | P value | Placebo<br>test P value |
| --- | --- | --- | --- | --- | --- | --- | --- |
| Q1 (lowest) | 1.5 | (0.9, 2.1) | < 0.001 | -- | -- | -- | -- |
| Q2 | 1.8 | (1.4, 2.2) | < 0.001 | 0.3 | (-0.1, 0.7) | 0.07 | 0.13 |
| Q3 | 2.0 | (1.6, 2.4) | < 0.001 | 0.5 | (0.1, 0.9) | 0.02 | 0.04 |
| Q4 | 2.1 | (1.5, 2.7) | < 0.001 | 0.6 | (0.2, 1.0) | 0.01 | 0.02 |
| Q5 (highest) | 2.4 | (1.4, 3.4) | < 0.001 | 0.9 | (0.1, 1.7) | 0.04 | 0.09 |

*Notes.* Model is a fully interacted OLS regression estimating the effects of physical distancing orders on neighborhoods in the lowest quintile for median household income, as well as the marginal effects of those policies on other income quintiles (interaction term). State, date, and lagged case count fixed effects are not reported. For placebo tests, the model was re-estimated 1000 times with simulated datasets in which policy exposure randomly reassigned across states, such that any estimated association was necessarily spurious. The placebo test *p* value reported is the proportion of iterations in which the placebo treatment produced a larger-magnitude *t* statistic than was estimated for the actual treatment. This metric is reported only for the estimated marginal effects relative to Q1.

**Figure 4:**
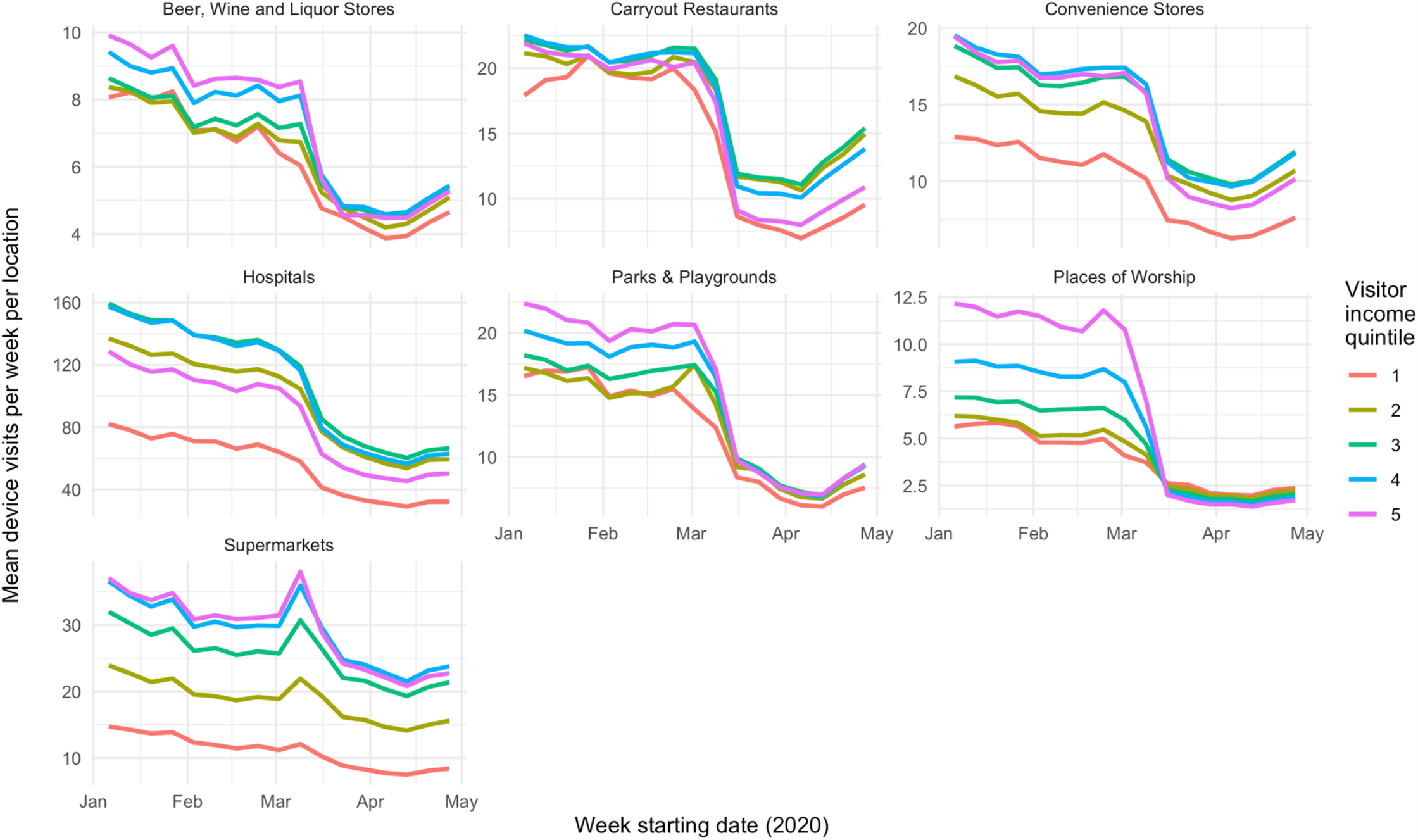
Visits to non-work locations. *Notes*: Calculated for locations in urban counties (population 100,000 and above) only. Visitor income is calculated for each point of interest based on visitor home census block group from January and February 2020. Median visitor income quintile is based on the median of household income values from visitors, weighted by the number of visits per BG.

The relationship between neighborhood income and changes in visits varied by category. For carryout restaurants and supermarkets, locations serving the lowest-income neighborhoods experienced the largest reductions in visits: visits to carryout restaurants declined by 57.6% (95% CI [55.8, 59.5]) for quintile 1, versus 54.4% (95% CI [53.5, 55.3]) for quintile 5, while visits to supermarkets declined by 39.0% (95% CI [37.3,40.7]) for quintile 1, compared to 33.6% (95% CI [32.0, 35.1]) for quintile 5. For three categories (beer, wine and liquor stores, convenience stores, and hospitals), the lowest-income neighborhoods reduced visits by a smaller percentage than the highest-income neighborhoods, but by a larger percentage than the other three income quintiles. Only for places of worship was there a gradient between income and reductions in visits. However, we found that visits in the post-period did not vary from weekday to weekends, suggesting that this pattern was not necessarily associated with visits for religious services. **Figure S2**.

**Fig. S2.**
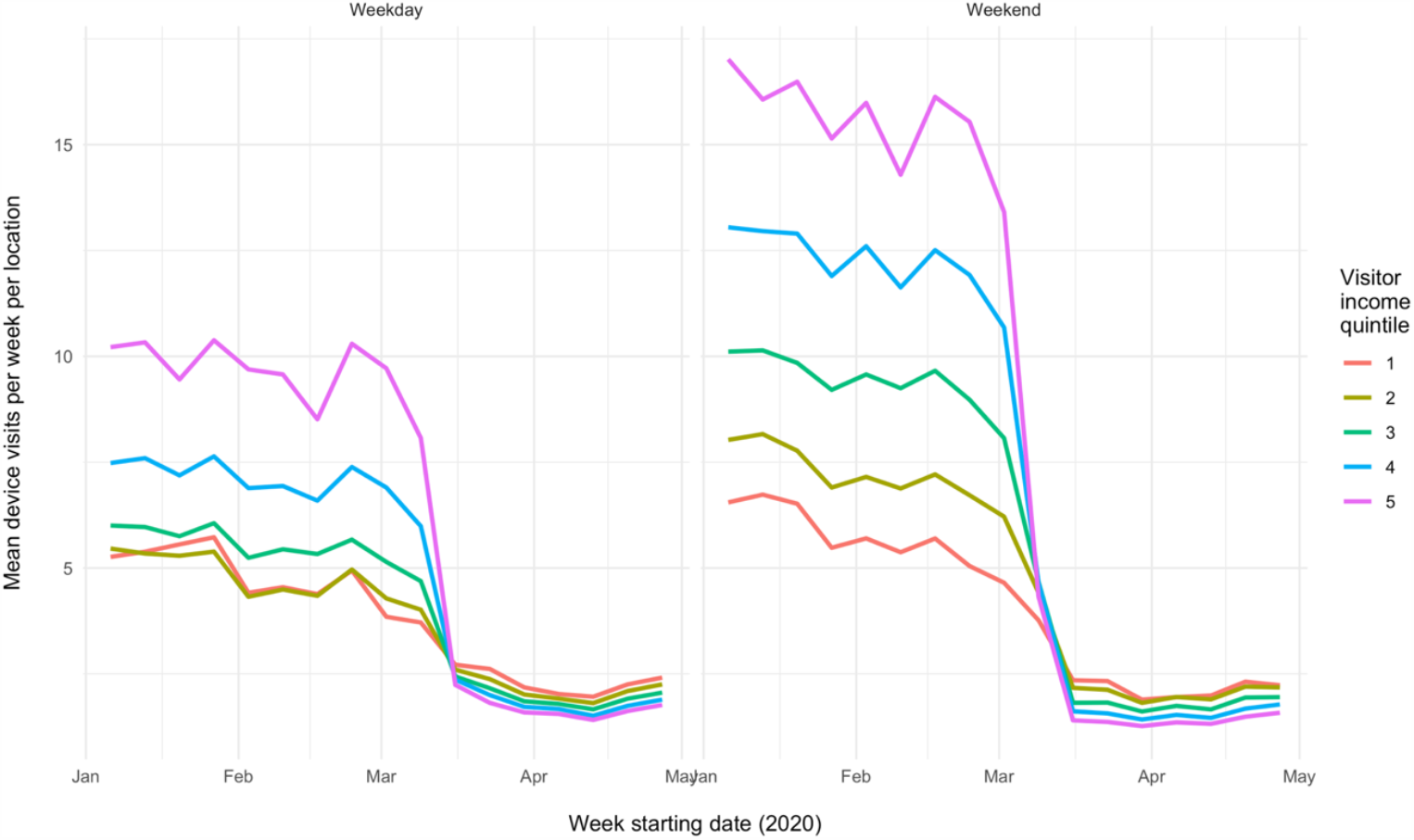
Visits to places of worship by weekday/weekend. *Notes:* Calculated for locations in urban counties (population 100,000 and above) only. Visitor income is calculated for each point of interest based on visitor home census block group (BG) from January and February 2020. Median visitor income quintile is based on the median of household income values from visitors, weighted by the number of visits per BG.

### State physical distancing orders

Our main DiD model found that physical distancing orders increased days spent at home for each neighborhood income quintile. In the lowest-income quintile, implementation of these orders was associated with a 1.5 percentage point increase in the proportion of smartphone users staying home all day (95% CI [0.9, 2.1], *p* < 0.001). **Table 3**. Treatment was also associated with *additional* increases in staying home for income quintiles 3, 4, and 5. In the highest neighborhood income quintile (*i*.*e*., quintile 5), physical distancing orders were associated with an additional 0.9 percentage point increase in staying home (95% CI [0.1, 1.7], *p* = 0.04). In other words, the estimated policy effects were 60% larger in the highest-income neighborhoods compared to the lowest-income neighborhoods. However, for all groups, the effects of state physical distancing orders were modest relative to the overall change in mobility observed during this period.

The placebo test validated statistical significance at *p* < 0.05 of the main effect (treatment effects in the lowest-income quintile) and for the marginal treatment effects for quintiles 3 and 4. For *treatment*quintile 5*, the placebo test-estimated *p* value was 0.09, *i*.*e*., 9% of the placebo runs reported an effect that appeared no less random than the estimated effect in our DiD model, even though the placebo design ensured that this effect was not real.

In the event study models, point-estimated increases in physical distancing were larger at higher income levels and generally statistically significant at *p* < 0.05 for at least one week after implementation for physical distancing orders. **Figure 5**.

**Figure 5:**
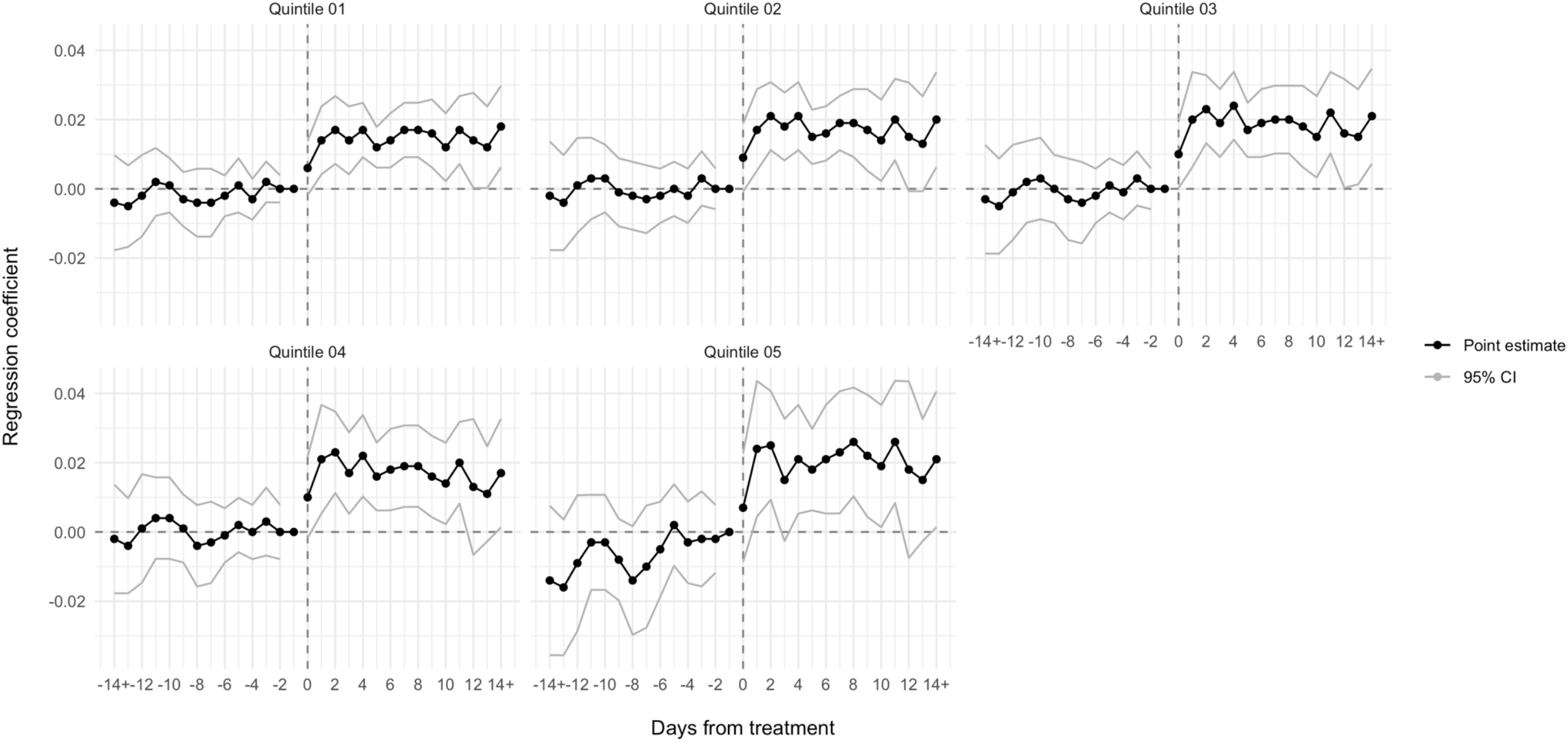
Event study linear regression analysis: effects of physical distancing orders on staying home all day. *Notes*. Each panel reports the result of an event study regression within a single income stratum. Models are identical to DiD models reported above, except that we replaced the binary policy indicator with binary indicators for living in intervention states in a series of one-day periods up to 14 days before and after policy changes. The reference group was being in a comparison state or being in an intervention state on the day before policy enactment (Day -1).

However, in the highest-income quintile, we found visible increases in distancing over the 14 days prior to the implementation of physical distancing orders. This pre-trend was not apparent in the other income quintiles.

## Discussion

Using national data, we found that communities at all income levels increased physical distancing in response to COVID-19. However, consistent with our first hypothesis, we found that lower-income communities increased physical distancing less than higher-income communities. We found evidence that working outside the home contributed to these differences in physical distancing, and no evidence that non-work activities outside the home contributed to these differences. Consistent with our second hypothesis, we found that state policies modestly increased physical distancing across all income levels, but increased them more at higher income levels.

Our findings indicate that income level is a strong determinant of whether individuals can stay home to protect themselves from COVID-19. Higher-income communities rapidly reduced the proportion of days that residents spent working outside the home, but our analysis suggests that lower-income communities could not. These findings are consistent with surveys indicating that while lower-income individuals wish to adopt physical distancing principles, they are unable to work from home.^8^ The lowest-income individuals might have experienced even smaller declines in working outside the home, had they not also lost work at a higher rate during the pandemic.^25^

In their non-work time, lower-income communities appear to have curtailed activities at similar rates as higher-income communities. In other words, it does not appear that non-work activities contributed to differences in physical distancing across income levels. Since physical distancing orders explicitly allowed essential workers to report for work, we find no evidence of higher *non-compliance* with these orders in low-income communities, contrary to an interpretation some prior studies have offered.^26^ Additionally, our findings may reflect differences in financial hardship across income levels. Larger reductions in visits to supermarkets and carryout restaurants serving low-income communities provides suggestive evidence that these communities experienced greater food insecurity than other communities. By contrast, larger reductions in visits to convenience stores in high-income communities may demonstrate those neighborhoods’ ability have convenience items delivered. Although media coverage has highlighted examples of non-compliance with physical distancing mandates within religious communities,^27^ our findings indicate that places of worship in low-income communities have likely fulfilled other functions during the COVID-19 pandemic (*e*.*g*., as food banks), consistent with their role in providing a “private safety net” for the poor.^28^

State physical distancing policies explicitly differentiated between workers who were expected to stay home and those who were expected to report to work. Our findings largely supported the hypothesis that such strategies disproportionately increased physical distancing in higher-income communities. Compared to the lowest-income neighborhoods, higher-income neighborhoods increased their days at home by larger amounts in response to physical distancing orders. The effect of these orders on the highest-income neighborhoods was somewhat less clear, particularly because it appeared that residents of these neighborhoods were steadily increasing days at home even before the orders went into effect.

### Limitations

This observational study is subject to several limitations. Although several research teams have used smartphone mobility data, including SafeGraph data, to study mobility trends, these data are novel and have not been validated against traditional data sources. Moreover, we lacked individual-level information on smartphone users, and therefore imputed user characteristics from BG data. Our sample was likely not representative of the overall population, since smartphone ownership varies, *e*.*g*., by age and income.^18^

We believe SafeGraph data track mobility trends more accurately than the absolute levels of the behaviors they measure. Trends in SafeGraph data appear to align with trends in data from similar smartphone location aggregation companies,^29^ and weekly trends in these data align with Gallup survey data on physical distancing practices.^30^ However, SafeGraph data could systematically over- or undercount the number of smartphone users staying home or going to work, in part because SafeGraph does not obtain data from every device at regular intervals through the day. Instead, the data represent locations from an irregularly timed sample of timepoints for each device throughout the day. As a result, there are periods in which a device is assumed to be at its last known location. We do not believe these errors would be correlated with socioeconomic position, supporting their use for comparing time trends across income levels.

In our analysis of state policy effects, we did not compare combinations of physical distancing policies, since the variation in these strategies was too limited for the time period studied. These questions should be the focus of future research. New opportunities to study these effects will emerge as the COVID-19 pandemic continues and jurisdictions dynamically adjust their responses. Additionally, we did not account for the influence of local policies, such as stay-at-home orders and curfews that city and county governments issued. These policies could potentially explain some of the physical distancing trends that state policies did not.

## Conclusions

The rapid inversion in the relationship between mobility and income during the COVID-19 pandemic illustrates how higher socioeconomic position affords greater opportunity to achieve good health. Staying home regularly was not an entrenched practice among higher-income individuals prior to COVID-19. On the contrary, spending days entirely at home was associated with worse health outcomes due to, *e*.*g*., physical inactivity,^31,32^ social isolation, and less utilization of healthcare. During the COVID-19 crisis, however, staying at home became a health seeking behavior. Although lower-income individuals had the knowledge and motivation to avoid exposure to COVID-19, as their reductions in non-work activities suggest, they were less able to stop reporting to work outside the home. Public policy reinforced these differences across income levels.

Financial barriers to physical distancing have likely contributed to a range of disparities in COVID-19 outcomes. Although governments have not published outcomes data by patient income level, outbreaks have been severe in U.S. cities, such as New Orleans and Detroit, with especially high poverty rates. In Massachusetts, as of May 20, the highest case counts per capita were found in Chelsea, Brockton, Lawrence, and other cities with high poverty rates.^33^ Moreover, since race, place, and poverty are closely interrelated,^34^ income-related disparities likely contribute to disproportionately high mortality rates for COVID-19 among African-Americans compared to other racial groups.^35,36^ Connections among communities may matter as well: for instance, Jung et al found a U-shaped relationship between county-level poverty and COVID-19 incidence, but only in high-density areas where high- and low-income residents might be most likely to cross paths.^17^

Our findings indicate that states must focus more on measures that enable lower-income residents to protect themselves through physical distancing. Policy options include restricting evictions, banning utility shut-offs, making unemployment insurance more readily available, and mandating paid sick leave.^9^ While these measures have not been adopted as widely as stay-at-home orders and non-essential business closures, they appear necessary to a more equitable COVID-19 response.

## Data Availability

All data are publicly available, except for SafeGraph smartphone mobility data. Access to these data for research purposes can be requested at the link provided.

https://www.safegraph.com/covid-19-data-consortium

## Acknowledgments

The authors thank SafeGraph for donating smartphone mobility data for this research study.

## Notes

### Competing Interest Statement

The authors have declared no competing interest.

### Funding Statement

No external funding was received for this research.

### Author Declarations

Since the mobility data were anonymized and other data were publicly available, this study was exempted from institutional review board review as non-human subjects research.

## References

1. Courtemanche, C., Garuccio, J., Le, A., Pinkston, J. & Yelowitz, A. Strong social distancing measures in the United States reduced the COVID-19 growth rate. Health Aff. (2020). doi:10.1377/hlthaff.2020.00608.

2. Ferguson, N. M. et al. Report 9: Impact of non-pharmaceutical interventions (NPIs) to reduce COVID19 mortality and healthcare demand. London: Imperial College London; 2020 Mar 16. Available at https://www.imperial.ac.uk/media/imperial-college/medicine/sph/ide/gida-fellowships/Imperial-College-COVID19-NPI-modelling-16-03-2020.pdf.

3. Chen, J. T. & Krieger, N. Revealing the unequal burden of COVID-19 by income, race/ethnicity, and household crowding: US county vs. ZIP code analyses. Preprint at https://cdn1.sph.harvard.edu/wp-content/uploads/sites/1266/2020/04/HCPDS_Volume-19_No_1_20_covid19_RevealingUnequalBurden_HCPDSWorkingPaper_04212020-1.pdf.

4. Chen, J. T., Waterman, P. D. & Krieger, N. COVID-19 and the unequal surge in mortality rates in Massachusetts, by city/town and ZIP code measures of poverty, household crowding, race/ethnicity,and racialized economic segregation. Preprint at: https://cdn1.sph.harvard.edu/wp-content/uploads/sites/1266/2020/05/20_jtc_pdw_nk_COVID19_MA-excess-mortality_text_tables_figures_final_0509_with-cover-1.pdf.

5. Gross, C. P. et al. Racial and Ethnic Disparities in Population Level Covid-19 Mortality. (2020). Preprint at: https://www.medrxiv.org/content/10.1101/2020.05.07.20094250v1.

6. Chowkwanyun, M. & Reed, A. L. Racial health disparities and Covid-19 — caution and context. N. Engl. J. Med. (2020). doi:10.1056/NEJMp2012910.

7. Williams, D. R. & Collins, C. Racial residential segregation: A fundamental cause of racial disparities in health. Public Health Rep. 116, 404–416 (2001).

8. Atchison, C. J. et al. Perceptions and behavioural responses of the general public during the COVID-19 pandemic: A cross-sectional survey of UK adults. Preprint at: https://www.medrxiv.org/content/10.1101/2020.04.01.20050039v1.

9. Raifman, J. et al. COVID-19 US State Policy Database. www.tinyurl.com/statepolicies (2020).

10. Garfield, R., Rae, M., Claxton, G. & Orgera K. Double Jeopardy: Low Wage Workers at Risk for Health and Financial Implications of COVID-19. Kaiser Family Foundation. https://www.kff.org/coronavirus-covid-19/issue-brief/double-jeopardy-low-wage-workers-at-risk-for-health-and-financial-implications-of-covid-19/ (2020).

11. Blau, F.D., Koebe, J. & Meyerhofer P.A. Essential and Frontline Workers in the COVID-19 Crisis. Econofact. https://econofact.org/essential-and-frontline-workers-in-the-covid-19-crisis (2020).

12. Cramer, R., O’Brien, R., Cooper, D. & Luengo-Prado, M. A Penny Saved is Mobility Earned. Washington, DC: Pew Charitable Trusts (2009).

13. Gupta, S. et al. Tracking public and private response to the Covid-19 epidemic. NBER Working Paper Series 27027 (2020). https://www.nber.org/papers/w27027.pdf.

14. Painter, M. & Qiu, T. Political beliefs affect compliance with COVID-19 social distancing orders. Preprint at: https://papers.ssrn.com/sol3/papers.cfm?abstract_id=3569098.

15. Andersen, M. Early evidence on social distancing in response to COVID-19 in the United States. Preprint at: https://papers.ssrn.com/sol3/papers.cfm?abstract_id=3569368.

16. Lasry, A. et al. Timing of community mitigation and changes in reported COVID-19 and community mobility ― Four U.S. metropolitan areas, February 26–April 1, 2020. MMWR. Morb. Mortal. Wkly. Rep. 69, (2020).

17. Jung, B. J., Manley, J. & Shrestha, V. Coronavirus infections and deaths by poverty status : Time trends and patterns. Preprint at: https://ideas.repec.org/p/tow/wpaper/2020-03.html.

18. Pew Research Center. Demographics of Mobile Device Ownership and Adoption in the United States. https://www.pewresearch.org/internet/fact-sheet/mobile/.

19. U.S. Bureau of Labor Statistics. Employment status of the civilian population by sex and age. https://www.bls.gov/news.release/empsit.t01.htm.

20. Ingram D.D. & Franco S.J. 2013 NCHS urban–rural classification scheme for counties. National Center for Health Statistics. Vital Health Stat 2:166 (2014).

21. The New York Times. An ongoing repository of data on coronavirus cases and deaths in the U.S. https://github.com/nytimes/covid-19-data.

22. Wing, C., Simon, K. & Bello-Gomez, R. A. Designing difference in difference studies: Best practices for public health policy research. Annu. Rev. Public Heal. 39, 453–469 (2018).

23. Bertrand, M., Duflo, E. & Mullainathan, S. How much should we trust differences-in-differences estimates? Q. J. Econ. 119, 249–275 (2004).

24. Chen, K., Cheng, Y., Berkout, O. & Lindhiem, O. Analyzing proportion scores as outcomes for prevention trials: a statistical primer. Prev. Sci. 18, 312–321 (2017).

25. Hrynowski, Z. COVID-19 Disrupts 30% of Americans’ Jobs or Finances. Gallup. https://news.gallup.com/poll/309299/covid-disrupts-americans-jobs-finances.aspx.

26. Wright, A. L., Sonin, K., Driscoll, J. & Wilson, J. Poverty and economic dislocation reduce compliance with COVID-19 shelter-in-place protocols. Preprint at: https://papers.ssrn.com/sol3/papers.cfm?abstract_id=3573637.

27. Stack, L. & Schweber, N. Defying virus rules, large Hasidic Jewish weddings held in Brooklyn. The New York Times March 17, 2020.

28. Edin, K. & Lein, L. The private safety net: The role of charitable organizations in the lives of the poor. Housing Policy Debate 9, 541–73 (1998).

29. Klein, B. et al. Assessing changes in commuting and individual mobility in major metropolitan areas in the United States during the COVID-19 outbreak. Preprint at: https://uploads-ssl.webflow.com/5c9104426f6f88ac129ef3d2/5e8374ee75221201609ab586_Assessing_mobility_changes_in_the_United_States_during_the_COVID_19_outbreak.pdf.

30. Kluch, S. The Compliance Curve: Will People Stay Home Much Longer? Gallup. https://news.gallup.com/opinion/gallup/309491/compliance-curve-americans-stay-home-covid.aspx.

31. Dewulf, B. et al. Associations between time spent in green areas and physical activity among late middle-aged adults. Geospat. Health 11, 225–232 (2016).

32. Simonsick, E. M., Guralnik, J. M., Volpato, S., Balfour, J. & Fried, L. P. Just get out the door! Importance of walking outside the home for maintaining mobility: Findings from the Women’s Health and Aging Study. J. Am. Geriatr. Soc. 53, 198–203 (2005).

33. Massachusetts Department of Public Health. COVID-19 Cases, Quarantine and Monitoring. https://www.mass.gov/info-details/covid-19-cases-quarantine-and-monitoring.

34. Tung, E. L., Cagney, K. A., Peek, M. E. & Chin, M. H. Spatial context and health inequity: Reconfiguring race, place, and poverty. J. Urban Heal. 94, 757–763 (2017).

35. Reyes, C., Husain, N., Gutowski, C., St. Clair, S. & Pratt. G. Chicago’s coronavirus disparity: Black Chicagoans are dying at nearly six times the rate of white residents, data show. Chicago Tribune Apr. 7, 2020.

36. Thebault, R., Tran A.B. & Williams, V. African Americans are at higher risk of death from coronavirus. The Washington Post Apr. 7, 2020.

